# Implementation Outcomes of a Web-Based Platform for Reporting and Monitoring Continuous Quality Improvement (CQI) Activities: A Mixed-Methods Evaluation

**DOI:** 10.64898/2026.03.31.26349902

**Authors:** Patience Komba, Georgia Simmonds, Elizabeth L. Dunbar, Kemar Bundy, Natalie Irving-Mattocks, Misti McDowell, Annette E. Ghee, Nancy Puttkammer

## Abstract

**Background:** Continuous Quality Improvement (CQI) is a core strategy for strengthening health systems, yet documentation and monitoring of CQI activities remain fragmented in many low- and middle-income country (LMIC) settings. In Jamaica, CQI has been institutionalized across priority programs, but it largely relies on paper-based tools and basic digital platforms that limit timely learning and oversight. To address these gaps, Jamaica’s Ministry of Health and Wellness (MOHW), in collaboration with the Caribbean Training and Education Centre for Health (C-TECH), adapted a web-based CQI application using a participatory, human-centered design approach.

**Methods:** We conducted a formative, convergent mixed-methods evaluation across 24 healthcare facilities to assess early-stage implementation of the CQI app. Guided by the Implementation Outcomes Framework, we examined acceptability, adoption, appropriateness, and feasibility. Quantitative data were collected through a structured survey of healthcare workers (n=43), and qualitative data were gathered through five focus group discussions (n=33) and three key informant interviews with CQI leads. Survey data were summarized descriptively, and qualitative data were analyzed using rapid qualitative analysis. Findings were integrated using joint displays.

**Results:** Survey findings indicated moderate to high perceived acceptability and appropriateness of the CQI app, with 70% of participants reporting that it saved time and 67% noting that it aligned with facility goals. However, 19% reported never using it. Qualitative findings highlighted the app’s value for improving CQI documentation, visualizing trends, and supporting supervisory oversight. Key barriers to sustained adoption included inconsistent internet connectivity, limited follow-up training, unclear team roles, and challenges integrating app use into routine workflows. Leadership engagement and alignment with existing CQI structures emerged as critical enablers.

**Conclusion:** This formative evaluation suggests that a digitally enabled CQI platform can strengthen documentation and oversight of quality improvement activities in resource-constrained health systems when embedded within supportive organizational and infrastructural contexts. Addressing foundational system readiness, including leadership engagement, capacity-building, and workflow integration, will be essential to realizing the CQI app’s potential in Jamaica and similar LMIC settings.

## 1. Introduction

Continuous Quality Improvement (CQI) is an evidence-based, structured, and widely used approach that systematically drives incremental improvements in healthcare services through iterative, data-informed decision-making (1, 2). CQI focuses on continually assessing healthcare processes, identifying opportunities for improvement, and adapting interventions to effectively respond to evolving healthcare demands and patient needs (1, 3). In practice, CQI involves health facility teams selecting priority quality indicators, iteratively testing process changes through Plan-Do-Study-Act (PDSA) cycles, reviewing performance data, and scaling changes that improve the quality of care (4). Its effectiveness has been demonstrated across diverse healthcare domains, including HIV care, maternal and child health, and the management of non-communicable diseases (NCDs), with documented improvements in clinical outcomes, patient retention, and service quality (1, 5–7).

Despite these advancements, persistent challenges continue to constrain CQI’s full potential. Fragmented health information systems, characterized by paper-based documentation practices and reliance on basic digital tools such as Microsoft Excel or Word, compound other barriers to effective CQI implementation, including disparities in service quality and limited resources, particularly human resources for health. These documentation and system-level constraints lead to incomplete and non-standardized data, hindering timely monitoring, cross-site learning, and evaluation of CQI interventions (1, 8–12). As a result, successful CQI changes often remain localized and are difficult to scale or institutionalize.

Jamaica provides a relevant context for examining these challenges. The country’s healthcare system is organized into four Regional Health Authorities (RHAs)- Western (WRHA), Southern (SRHA), Northeast (NERHA), and Southeast (SERHA), operating under the oversight of the Ministry of Health and Wellness (MOHW). Over the past decade, Jamaica has made notable progress in institutionalizing CQI, particularly within HIV care and chronic disease programs. Since 2013, the University of Washington, in collaboration with the Caribbean Technical Assistance and Education Centre for Health (C-TECH) and with funding from the U.S. President’s Emergency Plan for AIDS Relief (PEPFAR) through Health Resources and Services Administration (HRSA), has supported the MOHW in implementing multiple iterations of the Jamaica Quality Improvement Collaborative (JaQIC). These initiatives applied the Institute for Healthcare Improvement’s Breakthrough Series (BTS) model to strengthen HIV service delivery (4, 13). Early phases of JaQIC focused on improving the uptake and reliability of CD4 and HIV viral load testing, on-time antiretroviral (ART) medication pickup, and fostering a culture of quality data use and data-driven decision-making (13). These collaboratives supported multidisciplinary facility teams in testing and scaling evidence-based changes through structured learning sessions and peer exchange (14).

In collaboration with the MOHW, C-TECH has further advanced CQI as a core strategy for improving health outcomes across Jamaica by embedding structured CQI methods into routine service delivery at over 28 HIV treatment sites nationwide (15). By implementing PDSA cycles, facilities receive coaching from trained quality improvement (QI) coaches and participate in regular in-person or virtual learning sessions to promote shared learning and capacity building (16).

Despite these advances, CQI documentation in Jamaica has remained largely paper-based or reliant on basic software such as Microsoft Excel and Word. As CQI activities expanded across facilities and programs, these platforms increasingly constrain the ability to track progress over time, retain institutional memory, and enable system-level monitoring and learning. Similar challenges have been documented across other low-middle-income countries (LMIC) health systems, where weak documentation practices hinder the evaluation, dissemination, and institutionalization of effective quality improvement interventions (1, 3, 12)

To address these challenges and align with Jamaica’s broader digital health transformation agenda, (17, 18), C-TECH and the MOHW adopted a web-based CQI application (CQI app) designed to support standardized documentation, real-time monitoring, and cross-site learning of CQI activities. The CQI app was originally developed by the University of Maryland, Baltimore’s Center for International Health Education and Biosecurity (CIHEB) HIV program in Tanzania and later adapted for use in other settings, including Kenya, Rwanda, Zambia, and Botswana [19].

Evidence from digital health implementation research underscores the importance of meaningfully engaging healthcare workers and addressing technological self-efficacy to support the adoption and sustained use of digital tools (14, 19, 20). Similarly, digital interventions that are grounded in local contexts and shaped by end users’ perspectives have been shown to be more acceptable, relevant, and sustainable (21). Guided by this evidence and recognizing that the CQI app was not originally designed for the Jamaican context, C-TECH and the MOHW, supported by the Digital Initiatives Group at I-TECH (DIGI), University of Washington, undertook a participatory, human-centered adaptation process of the CQI app. This approach focused on collaboratively assessing the app’s fit for purpose and identifying context-specific adaptations needed to align the tool with healthcare providers’ needs, capacities, and operational realities within the Jamaican health system (22).

We conducted an early formative evaluation to explore initial implementation outcomes of the CQI app and identify existing challenges to inform adaptation, adoption, and scale-up. The goal was to generate practical insights to support the successful integration of the app into routine practice and to strengthen the overall CQI process across Jamaica’s health system. Specifically, the evaluation sought to examine:

1. Acceptability of the CQI app among healthcare providers within Jamaican healthcare facilities
2. Extent of adoption of the CQI app among healthcare providers in Jamaica
3. Perceived appropriateness of the CQI app in the Jamaican health care system
4. Feasibility of implementing the CQI app within Jamaican healthcare settings

The evaluation was not designed to assess effectiveness or impact on clinical outcomes, but rather to generate implementation-relevant insights to inform ongoing adaptation, integration, and potential scale-up of the CQI app within Jamaica’s health system and similar LMIC contexts.

## 2. Methods

### Evaluation Setting

This evaluation was conducted in Jamaica and involved 24 healthcare facilities across the country’s four RHAs: WRHA, SRHA, NERHA, and SERHA. Participating facilities provide a range of health services, including HIV care and treatment, management of NCDs, and mental health services. All sites were engaged in CQI activities supported by C-TECH and the MOHW.

At each facility, CQI teams implemented QI processes guided by the PDSA framework. These activities included regular team meetings, performance monitoring to identify gaps in service delivery, root cause analysis, development and testing of change ideas, and systematic documentation of CQI efforts. CQI teams also participated in scheduled learning sessions that supported peer-to-peer learning, shared problem-solving, and capacity building. These learning sessions served as a forum for healthcare workers (HCWs) to present ongoing CQI initiatives and receive targeted didactic support to strengthen CQI implementation.

### The CQI app intervention

The CQI app is a web-based platform based on the PDSA framework that supports health facility teams in documenting, monitoring, and sharing their CQI activities. The platform was developed to address persistent challenges in CQI documentation and limited opportunities for team-based and peer learning of HIV care and treatment quality improvement programs.

The CQI app includes two primary modules. The facility module (Figure 1) enables frontline CQI teams to log projects, track indicators, and document PDSA cycles. The manager module (Figure 2) allows supervisors and CQI coaches to view dashboards, monitor trends across many facilities, and provide remote support. The platform supports visualization of CQI progress over time, facilitates real-time performance monitoring, and serves as a repository for CQI-related job aids, training materials, and guidelines.

**Figure 1:**
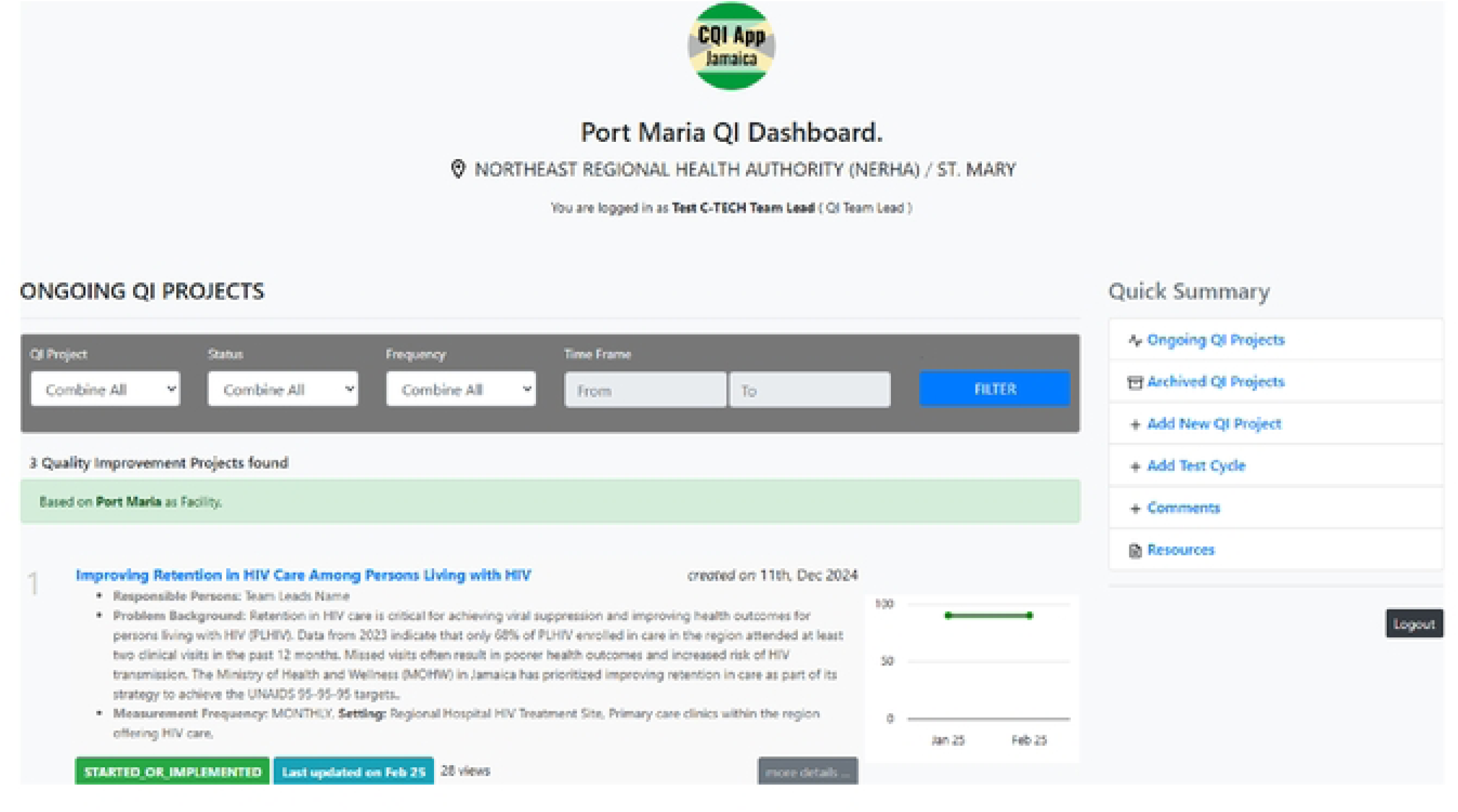
CQI app- Facility Module.

**Figure 2:**
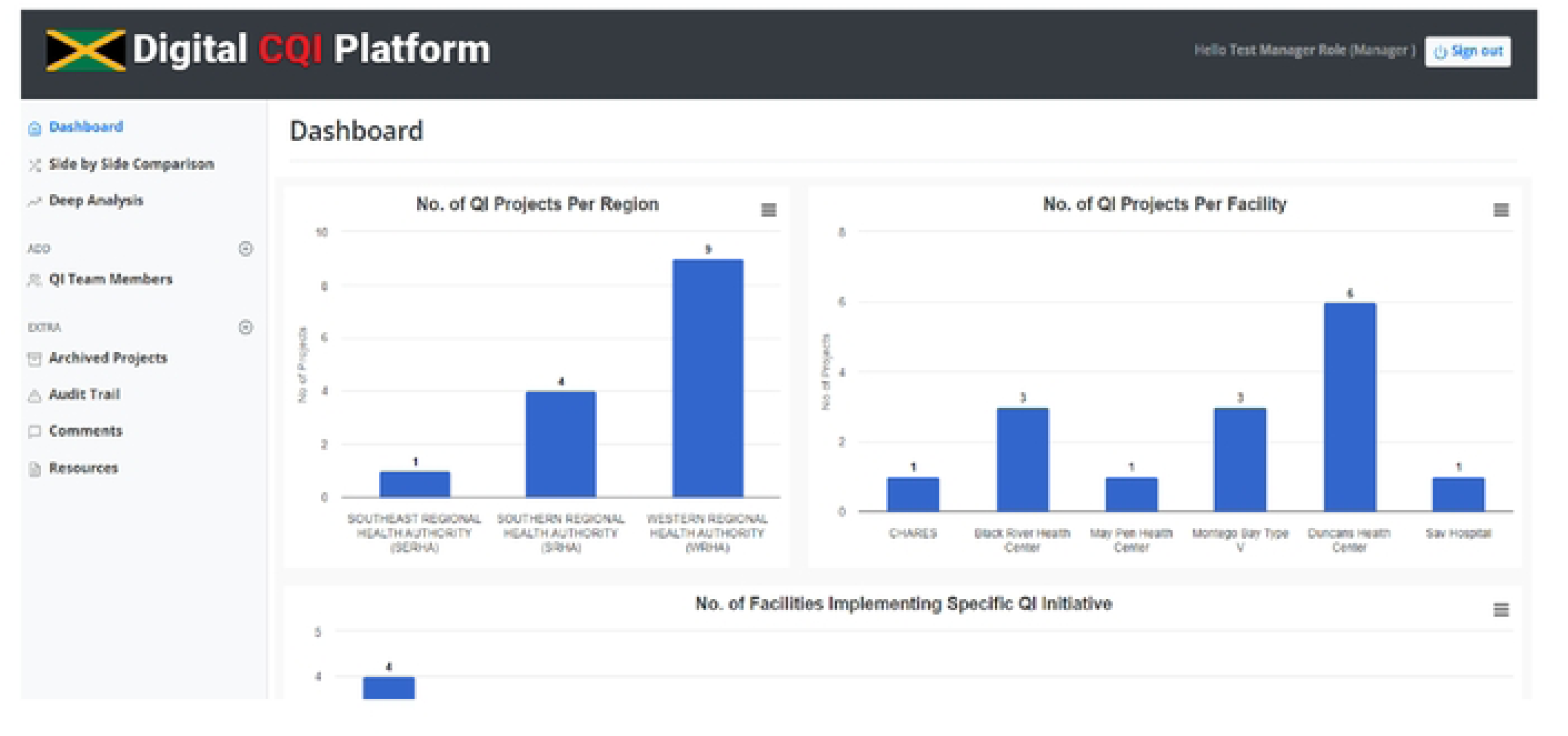
CQI app- Managers Module.

C-TECH became aware of the CQI app through publications and information shared on UMB-CIHEB’s programme website (23, 24). Given the similarity in CQI implementation contexts and health system constraints across these LMIC settings, C-TECH and the MOHW explored the CQI app as a potential solution to Jamaica’s CQI documentation and monitoring challenges. Initial engagement involved direct consultations with the UMB-CIHEB development team in Tanzania to understand the app’s functionality, implementation approach, and applicability to the Jamaican context. Following these consultations, C-TECH and the MOHW decided to adapt and pilot the CQI app in Jamaica.

The adaptation process followed a participatory, human-centered design approach and involved multiple co-design sessions with HCWs, C-TECH staff, and MOHW representatives. These sessions focused on identifying Jamaica-specific CQI documentation needs and workflow requirements. While the app’s core structure mirrored the standard PDSA-based CQI process, adaptations included configuring facility and regional identifiers, aligning terminology with Jamaican health system workflows, and tailoring user roles and reporting structures to reflect local CQI implementation practices.

Implementation of the CQI app in Jamaica included orientation sessions and training workshops for HCWs and CQI coaches, followed by iterative testing and refinement of the platform between July 2022 and November 2023. The app was introduced to facilities as a complementary tool to existing CQI processes, enabling teams to gradually integrate digital documentation into routine practice. The first facility-based CQI project was formally documented in the CQI app in December 2023.

### Evaluation Design

This evaluation employed a convergent mixed-methods design to assess early implementation of the CQI app across participating healthcare facilities. We collected quantitative data from a survey and qualitative data from Focus Group Discussions (FGDs) and Key Informant Interviews (KIIs) concurrently, analyzed them separately, and integrated them during interpretation to provide a comprehensive understanding of the CQI app implementation experiences (Figure 3). The evaluation was guided by the Implementation Outcomes Framework, focusing on four early-stage outcomes: acceptability, adoption, appropriateness, and feasibility. These outcomes were selected to align with the evaluation timing during the initial rollout of the CQI app, when later-stage outcomes such as penetration, fidelity, cost, and sustainability had not yet been sufficiently established for robust assessment (25).

**Figure 3:**
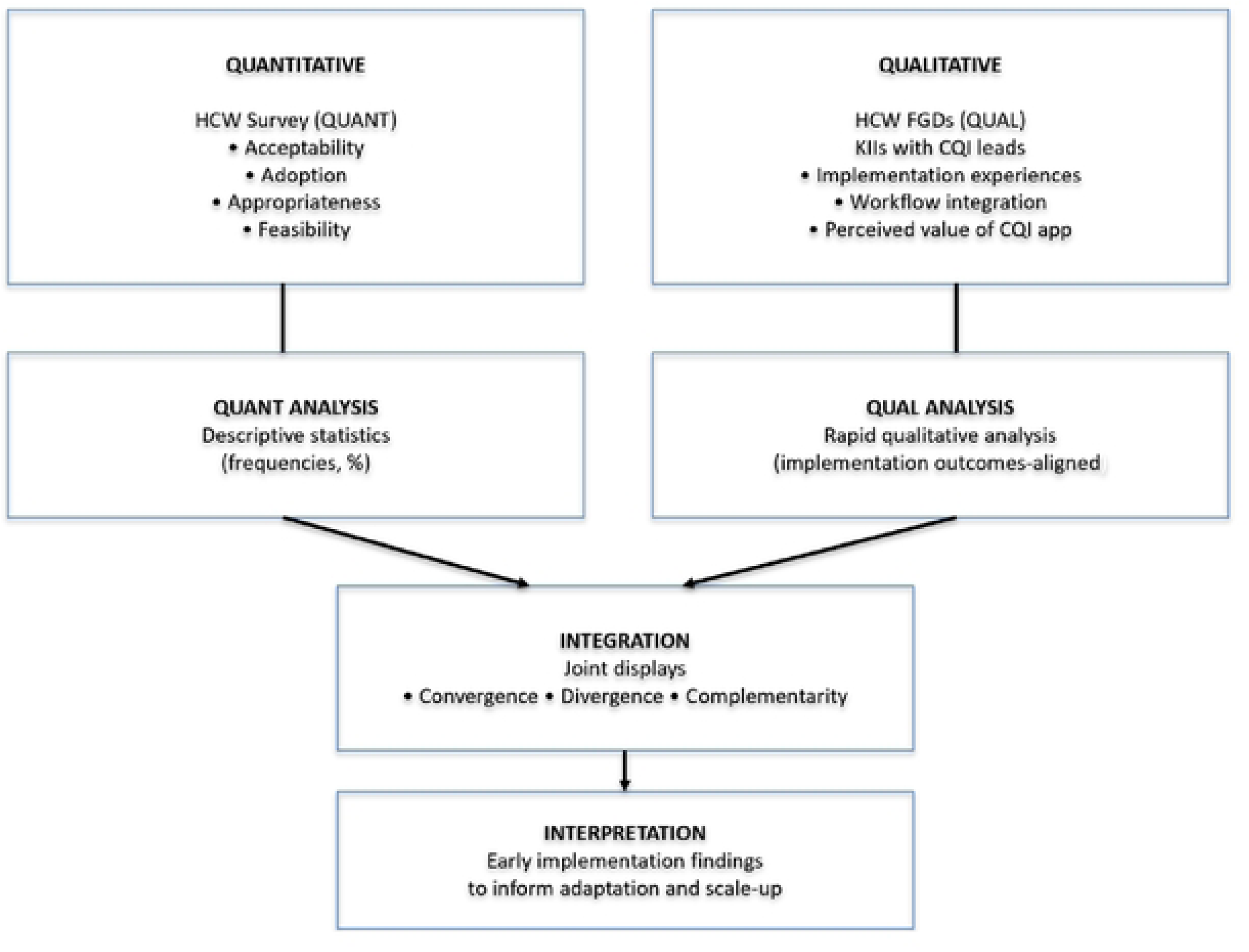
Convergent Mixed Methods Design- Adapted from Edmonds & Kennedy (2017)(26).

Quantitative data were collected through a structured survey to capture HCWs’ perceptions of the CQI app across the four implementation outcomes. Qualitative data were collected through FGDs and KIIs to explore contextual factors influencing implementation, including workflow integration, training experiences, and perceived value of the CQI app for routine CQI practice.

Findings from the quantitative and qualitative components were integrated using joint displays to enable side-by-side comparison and interpretation. This approach facilitated the identification of convergence, divergence, and complementarity among data sources, thereby strengthening the interpretation of early implementation findings.

#### Evaluation Population

Participants were HCWs from the 24 facilities across WRHA, SRHA, NERHA, and SERHA, supported by C-TECH and the Jamaica MOHW, collectively known as the CQI network.

### Recruitment of Participants

#### Human Subjects’ Protection

All participants provided verbal consent at the beginning of each FGD and KII. Evaluation procedures ensured that no personally identifiable information (PII) was collected and that confidentiality was maintained throughout the evaluation. Participants were assigned unique identifiers (e.g., A1, A2, B1, B2) and referred to using these codes during discussions.

Audio recordings were securely stored during the analysis phase and destroyed thereafter. Field notes and transcripts were anonymized and de-identified before analysis.

Invitations to participate in the survey were emailed and included a Participant Information Sheet (PIS) describing the purpose of the evaluation, voluntary participation, and anonymity, as well as a secure link to the online survey.

The Jamaica MOHW approved this evaluation as part of program activities funded by HRSA, which support the C-TECH-led HIV/AIDS care and treatment program. The University of Washington Human Subjects Division (HSD) also granted a human subject research exemption status, IRB ID: STUDY00020691.

##### Survey Participants

Survey respondents were HCWs who had previously received training in CQI methods and had been exposed to the CQI app during routine programme activities. Eligible participants were identified using CQI and CQI app training records maintained by C-TECH and MOHW. The survey was distributed to all HCWs listed in these training logs, regardless of whether they had begun using the CQI app at the time of data collection.

This inclusive approach was intentional, as the evaluation aimed to capture early perceptions of the CQI app across varying levels of exposure and use, including perspectives from HCWs who had not yet integrated the app into routine practice. The survey targeted diverse professional cadres involved in CQI implementation, including adherence counsellors, case managers, contact investigators, clinicians, health facility managers, psychologists, social workers, and data management staff such as data entrants and Treatment Care Support Officers (TCSOs). This approach ensured representation across key roles engaged in CQI activities at the facility level.

##### Participants for Focus Group Discussion and Key Informant Interviews

Participants for FGDs and KIIs were selected using criterion-based purposive sampling from attendees of semi-annual CQI learning sessions conducted between June and July 2024 as part of routine CQI programming (27). All attendees at these learning sessions were CQI team members or CQI coaches representing their respective facilities or regions.

Selection of participants for qualitative data collection was conducted jointly by C-TECH programme staff in consultation with MOHW CQI leads. HCWs who had previously received CQI app training, whether they had already used the CQI app, were invited to participate in FGDs. Eligibility criteria for FGDs included: (1) participation in CQI activities at the facility or regional level, (2) prior CQI training, and (3) exposure to CQI app training. No additional exclusion criteria were applied.

FGDs included 6–8 participants per group and were conducted immediately following the learning sessions to allow reflection on both the session content and participants’ ongoing CQI implementation experiences. Participants were selected to ensure representation across professional roles, facility and regional affiliations, and varying levels of CQI app use, including both users and non-users.

KIIs were conducted with QI coaches and regional managers responsible for CQI oversight. These KIIs were carried out during the same evaluation period as the learning sessions and FGDs. Eligibility criteria for KIIs included holding a formal CQI oversight role at the facility or regional level and prior engagement with CQI implementation processes. KIIs focused on system-level implementation dynamics, supervisory perspectives, and contextual factors influencing CQI app integration. While FGDs and KIIs were conducted in parallel during the evaluation period, insights from early FGDs informed the areas of emphasis in subsequent KIIs, consistent with a convergent mixed-methods evaluation design.

#### Evaluation Procedures

Quantitative Data Collection: We collected quantitative data using a Google Forms survey structured around four implementation outcomes: acceptability, adoption, appropriateness, and feasibility. The survey included five demographic questions and 19 Likert-scale items capturing HCWs’ perceptions and experiences with the CQI app. Each item used a five-point response scale ranging from strongly disagree to strongly agree. Survey items assessed perceived ease of use, usability, alignment with CQI goals, and feasibility of integrating the CQI app into existing workflows.

Examples of survey items by implementation outcome included:

(i) Acceptability: *“The CQI app is easy to use in my daily tasks.”*
(ii) Adoption: *“I would like to use the CQI app more frequently.”*
(iii) Appropriateness: *“The CQI app aligns well with our facility’s goals.”*
(iv) Feasibility: *“It is feasible to use the CQI app with the current level of IT support.”*

The full survey instrument is provided under supporting information.

We distributed the survey via email one week (26^th^ June 2024) prior to the FGDs and KIIs and allowed participants about one month to respond. Data collection closed after all qualitative activities were completed (31^st^ July 2024). Quantitative survey data were analysed as described in the data analysis section below.

#### Qualitative data collection

We collected qualitative data through five FGDs and three KIIs, all conducted in English, alongside scheduled CQI learning sessions. Each FGD included 6–8 participants and lasted approximately 45–60 minutes, while each KII included one participant and lasted approximately 30 minutes.

At the end of each learning session day, C-TECH programme staff invited eligible participants to take part in FGDs and moved them to a separate room, where they provided information about the evaluation and obtained verbal consent. An evaluation specialist from the Digital Initiatives Group at I-TECH, University of Washington, travelled to Jamaica and led the qualitative data collection activities. Three FGDs were conducted in person during the learning sessions. Due to unforeseen logistical disruptions during the evaluation period, the remaining two FGDs and all KIIs were conducted with the interviewer joining remotely via Zoom, while participants remained on site at the learning session venues. This change in interviewer modality did not alter the discussion guides, participant composition, or data collection procedures.

We audio-recorded all FGDs and KIIs and transcribed them verbatim. Qualitative data from FGDs and KIIs were analysed using the approach described in the Data Analysis section below.

##### Quantitative Analysis

We exported survey data from Google Forms to Microsoft Excel for initial cleaning and subsequently analysed it using Stata version 17. We calculated descriptive statistics, including frequencies and percentages, to summarize participant demographics and survey responses. We analysed Likert-scale responses across the four implementation outcomes to assess HCWs’ perceptions of the CQI app.

##### Qualitative Analysis

We analysed qualitative data using rapid qualitative analysis (RQA), a pragmatic approach commonly applied in implementation research to generate timely, actionable insights through structured and iterative data review (28). RQA aligned with the evaluation’s objective of informing programme improvement during early implementation. The approach combines deductive analysis of predefined constructs with inductive identification of context-specific themes. It is well documented for its efficiency and for generating actionable findings in time-and resource-constrained contexts while maintaining analytical rigor (29, 30).

We used structured data summary templates to systematically extract and organize information from FGDs and KIIs across the four implementation outcomes. These templates supported the identification of key themes, illustrative quotes, and relevant contextual factors. Because all FGDs and KIIs were audio-recorded and transcribed verbatim, we reviewed the rapid summaries alongside full transcripts to enhance analytical completeness and validity.

Figure 4 shows key phases of the adaptation, implementation, and evaluation of the CQI app.

**Figure 4:**
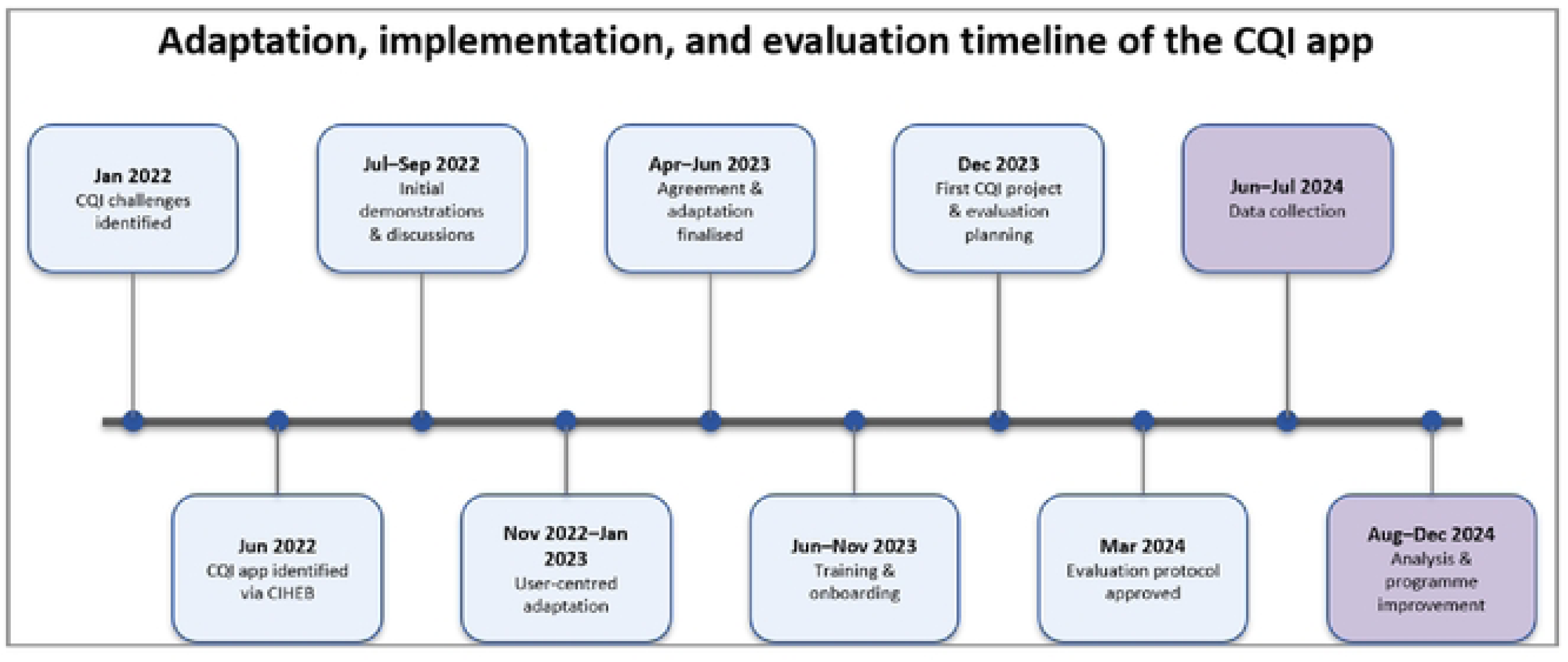
Adaptation, Implementation, and Evaluation Timeline of the CQI app (January 2022 to December 2024)

##### Integration of Findings

We analyzed the quantitative and qualitative data separately to maintain the methodological integrity and unique strengths of each dataset. The findings were then integrated using joint display, a mixed-methods strategy that supports side-by-side interpretation of quantitative and qualitative findings (31). This approach facilitated triangulation by identifying areas of convergence (where findings aligned), divergence (where responses differed), and complementarity (where qualitative insights added explanatory depth to survey results), while capturing participants’ voices and providing contextual understanding (32).

## 3. Results

### Descriptive summary of participants’ characteristics

*Table 1* presents the characteristics of participants in the evaluation’s quantitative and qualitative components. For the quantitative component, 43 healthcare workers completed the survey, yielding a 65% response rate from the 66 individuals invited. For the qualitative component, five FGDs involving 33 participants were conducted. In addition, KIIs with three senior healthcare personnel were conducted to assess their roles in overseeing CQI implementation.

**Table 1:** Participants’ Characteristics for Qualitative and Quantitative Evaluation Components.

| *Participants Characteristics* | *Quantitative Data*<br>*(Survey)*<br>*n = 43 (%)* | *Qualitative Data*<br>*(FGD)*<br>*n = 33 (%)* |
| --- | --- | --- |
| **Regions Represented** |  |  |

| ***Participants Characteristics*** | ***Quantitative Data***<br>***(Survey)***<br>***n = 43 (%)*** | ***Qualitative Data***<br>***(FGD)***<br>***n = 33 (%)*** |
| --- | --- | --- |
| WRHA | 11 (26%) | 9(28%) |
| SERHA | 14 (32%) | 12(37%) |
| SRHA | 10 (23%) | 6(19%) |
| NERHA | 8 (19%) | 5(16%) |
| **Years of involvement with CQI** |  |  |
| <1 year | 16 (37%) | <1 year: 10 (30%) |
| 1–2 years | 10 (23%) | 1–2 years: 8 (24%) |
| 3–5 years | 7 (16%) | 3–5 years: 5 (15%) |
| 6–10 years | 6 (14%) | 6–10 years: 3 (9%) |
| >10 years | 4 (9%) | >10 years: 7 (21%) |
| **Professional Cadres** |  |  |
| TCSOs/Data Entrants | 18 (42%) | 10 (30%) |
| Doctors/Clinicians | 6 (14%) | 8 (24%) |
| Social Workers | 6 (14%) | 3 (9%) |
| Contact Investigators | 5 (12%) | 2 (6%) |
| Adherence Counselors | 4 (9%) | 6 (18%) |
| Case Managers | 2 (5%) | - |
| Psychologist | 1 (3%) | - |
| Health Manager | 1 (3%) | - |
| QI Coaches/Leads | - | 2 (6%) |
| Data Manager | - | 1 (3%) |

**Key:** WRHA: Western Regional Health Authority, SERHA: Southeast Regional Health Authority, SRHA: Southern Regional Health Authority, NERHA: Northeast Regional Health Authority, TCSO: Treatment Care Support Officer, QI: Quality Improvement.

#### Experience with Healthcare, CQI, and the CQI app

Most survey participants (65%) had extensive healthcare experience, with 37% reporting 6–10 years and 28% more than 10 years (Table 2). Smaller proportions reported 3–5 years (14%), 1–2 years (12%), and less than 1 year (9%). Participants varied in their familiarity with CQI frameworks: 47% had less than 1 year of CQI experience, 26% had 1–2 years, 14% had 3–5 years, 9% had 6–10 years, and 5% reported more than 10 years. Regarding CQI app usage, 67% of participants used it 1–2 times, 12% used it monthly, 2% used it weekly, and 19% reported never using it.

**Table 2:**
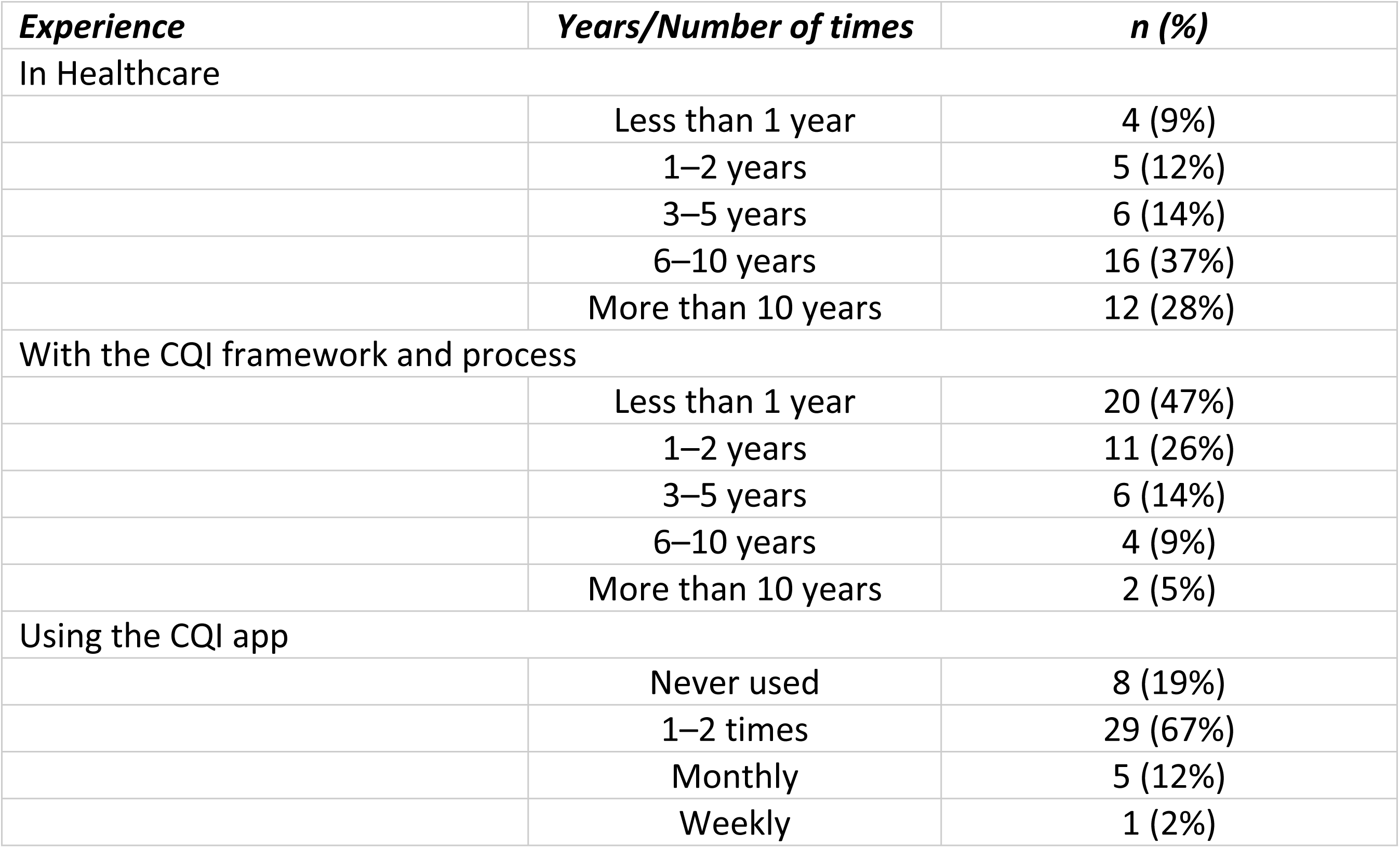
Survey participants’ Experience in Healthcare, Familiarity with CQI Frameworks, and CQI app Usage (n=43)

| *Experience* | *Years/Number of times* | *n (%)* |
| --- | --- | --- |
| In Healthcare |  |  |
|  | Less than 1 year | 4 (9%) |
|  | 1–2 years | 5 (12%) |
|  | 3–5 years | 6 (14%) |
|  | 6–10 years | 16 (37%) |
|  | More than 10 years | 12 (28%) |
| With the CQI framework and process |  |  |
|  | Less than 1 year | 20 (47%) |
|  | 1–2 years | 11 (26%) |
|  | 3–5 years | 6 (14%) |
|  | 6–10 years | 4 (9%) |
|  | More than 10 years | 2 (5%) |
| Using the CQI app |  |  |
|  | Never used | 8 (19%) |
|  | 1–2 times | 29 (67%) |
|  | Monthly | 5 (12%) |
|  | Weekly | 1 (2%) |

### Survey Results by Implementation Outcome

#### Acceptability

Participants expressed mixed perceptions of the CQI app’s acceptability (Table 3). Just over half of respondents reported overall satisfaction with the app, while a substantial proportion expressed neutral views, suggesting tentative acceptance during the early implementation phase. More than half of the respondents found the app pleasant to use and did not perceive it as unnecessarily complex. A similar proportion agreed that the app met their CQI documentation needs. However, interest in more frequent use was moderate, indicating that acceptability did not consistently translate into sustained engagement.

**Table 3.**
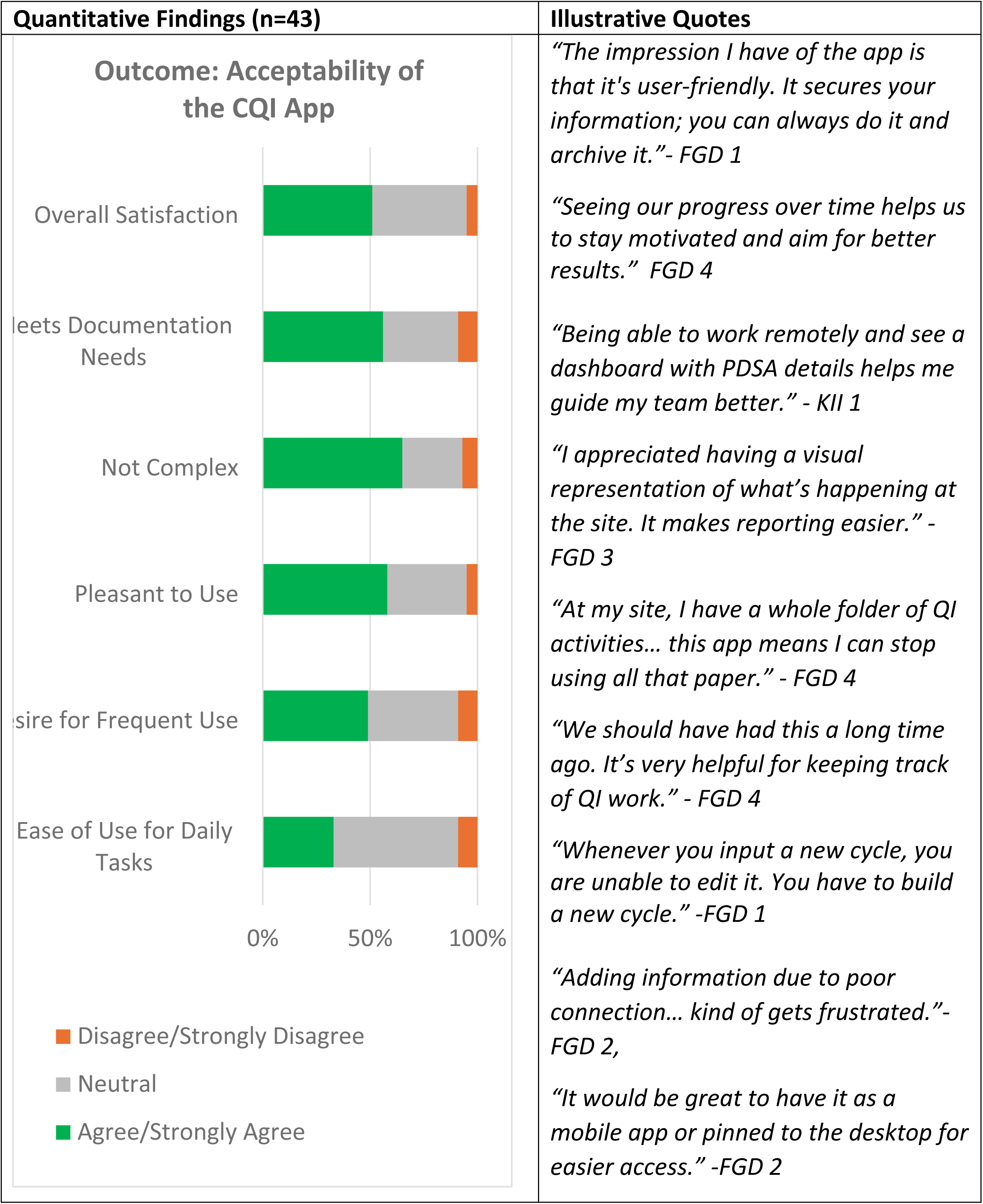
Joint Display of Mixed Methods Findings on the Acceptability of the CQI App.

Qualitative findings helped contextualize these patterns. Participants highlighted positive features such as the user-friendly dashboard, project archiving, and the ability to track CQI trends over time, which supported reporting and team engagement. At the same time, participants described internet instability, password reset challenges, and difficulties with updating CQI project entries as common barriers undermining usability. Several participants reported limited use of the CQI app following training, citing workload constraints and limited follow-up support.

Participants suggested several improvements to enhance acceptability, including simplifying the interface, developing a mobile version, adding autosaving functionality, and incorporating reminders or prompts (job aids) to support ongoing use.

#### Adoption

Participants reported generally positive but uneven perceptions of the CQI app’s adoption (Table 4). Most respondents indicated willingness to recommend the app and perceived it as time-saving, and a majority felt it supported the sharing of CQI data. However, fewer participants perceived clear improvements in their own CQI work, suggesting that perceived value did not consistently translate into deeper integration into routine practice.

**Table 4.**
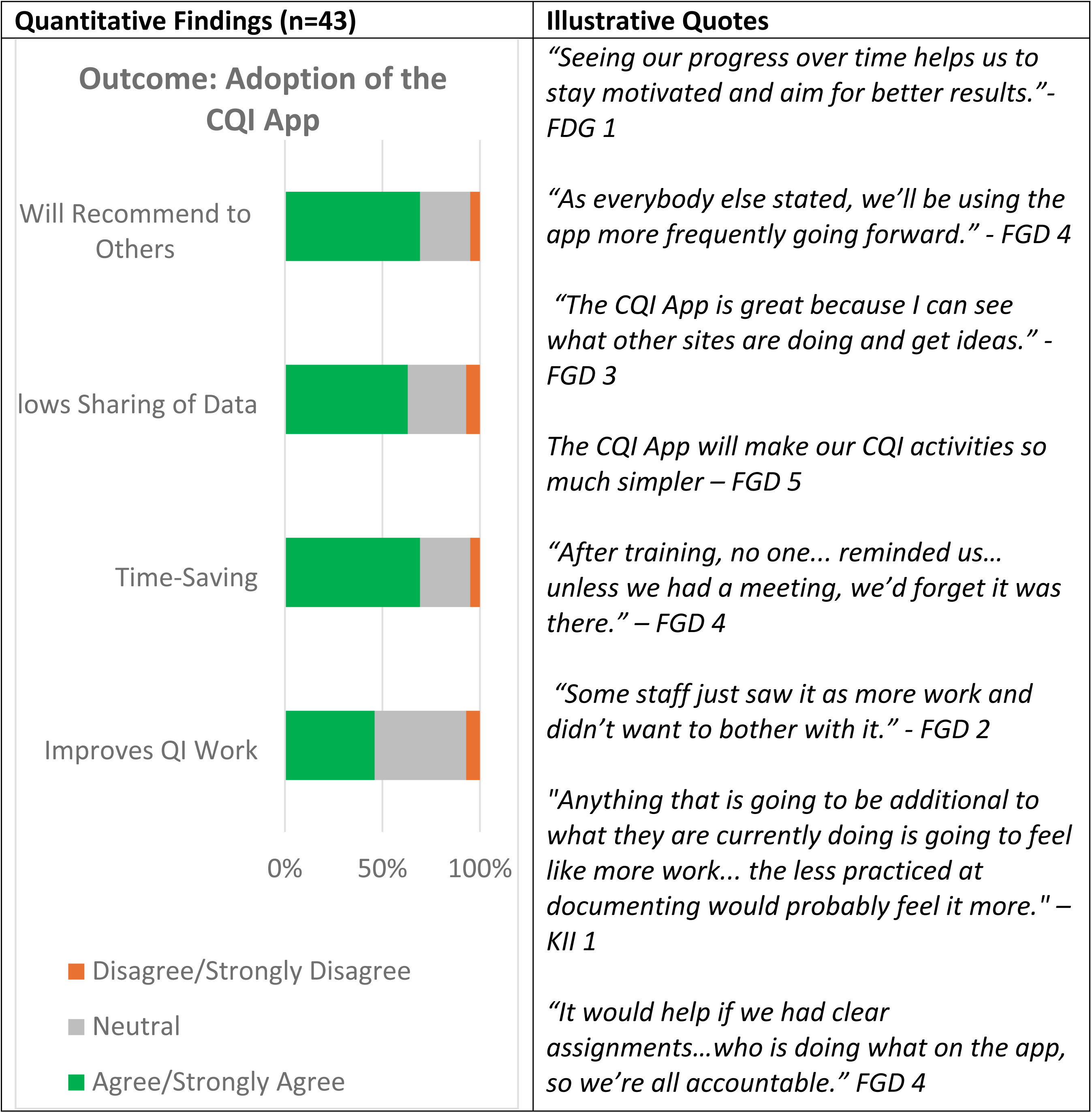
Joint Display Mixed Methods Findings on the Adoption of the CQI App.

Qualitative findings provided insight into these patterns. Participants highlighted the app’s ability to store and retrieve CQI documentation, monitor progress remotely, and facilitate collaboration as features that improved efficiency and saved time. At the same time, participants described barriers to adoption, including limited follow-up after initial training, unclear roles for app use, and competing workload demands. Participants suggested expanding team-wide training, clarifying user responsibilities, and integrating CQI app tasks more explicitly into routine workflows might increase the app’s adoption.

#### Appropriateness

Participants generally perceived the CQI app as appropriate for their work and aligned with facility goals (Table 5). Most respondents indicated that the app was suitable for their tasks and relevant to their roles, and that it supported documentation standards. At the same time, responses suggested more mixed views regarding whether the use of the CQI app reduced workload or avoided adding burden.

**Table 5.**
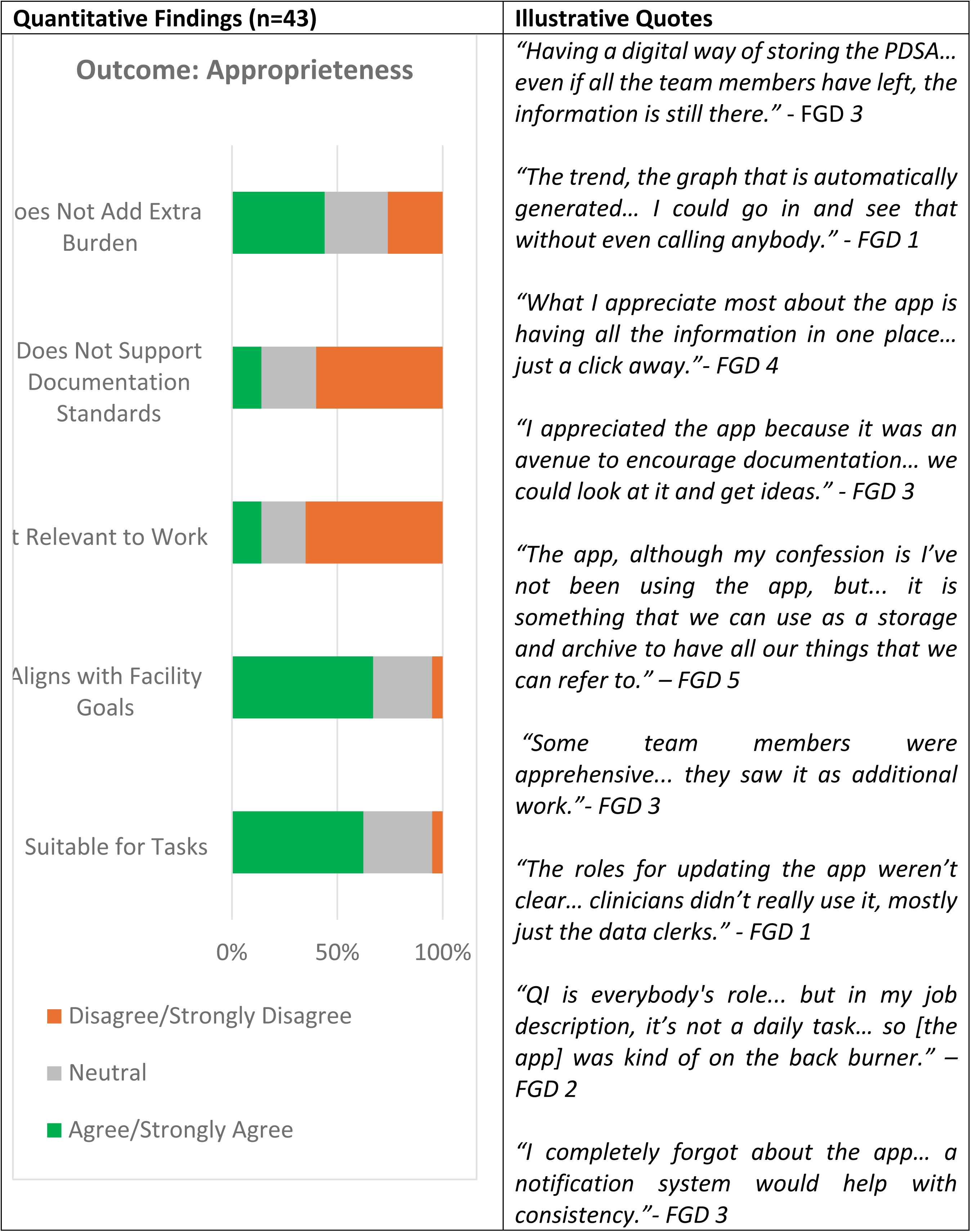
Joint Display of Mixed Methods Findings on the Appropriateness of the CQI App. Note:Survey items assessing appropriateness included both positive and negatively framed statements. For negatively worded items, disagreement reflects a positive appropriateness outcome.

Participants described several features that supported the app’s appropriateness, including the ability to view and track CQI data over time, auto-generated graphs, and centralized dashboards that made information easier to access. Several participants noted that the app aligned well with documentation expectations and supported both clinical and managerial oversight. QI coaches emphasized the app’s value for consolidating facility-level CQI data and maintaining institutional memory through its archiving functionality.

Participants also described challenges related to workflow integration. Many viewed the CQI app as an additional task rather than a routine component of daily work. Responsibilities for app updates were often unclear, with clinicians and psychosocial staff frequently relying on data clerks to enter data. In some facilities, the absence of reminders or clearly designated roles limited consistent use. Participants suggested that clearer role definitions, routine expectations for updates, and ongoing training tailored to different team members could strengthen alignment with existing workflows.

#### Feasibility

Participants expressed mixed views regarding the feasibility of the CQI app (Table 6). A majority perceived the app as feasible to use with the existing level of IT support, and most did not report frequent technical issues. However, responses were less definitive regarding the availability of resources to support routine use and the consistency of updates and maintenance, with a substantial proportion of respondents indicating neutral positions on these items.

**Table 6.**
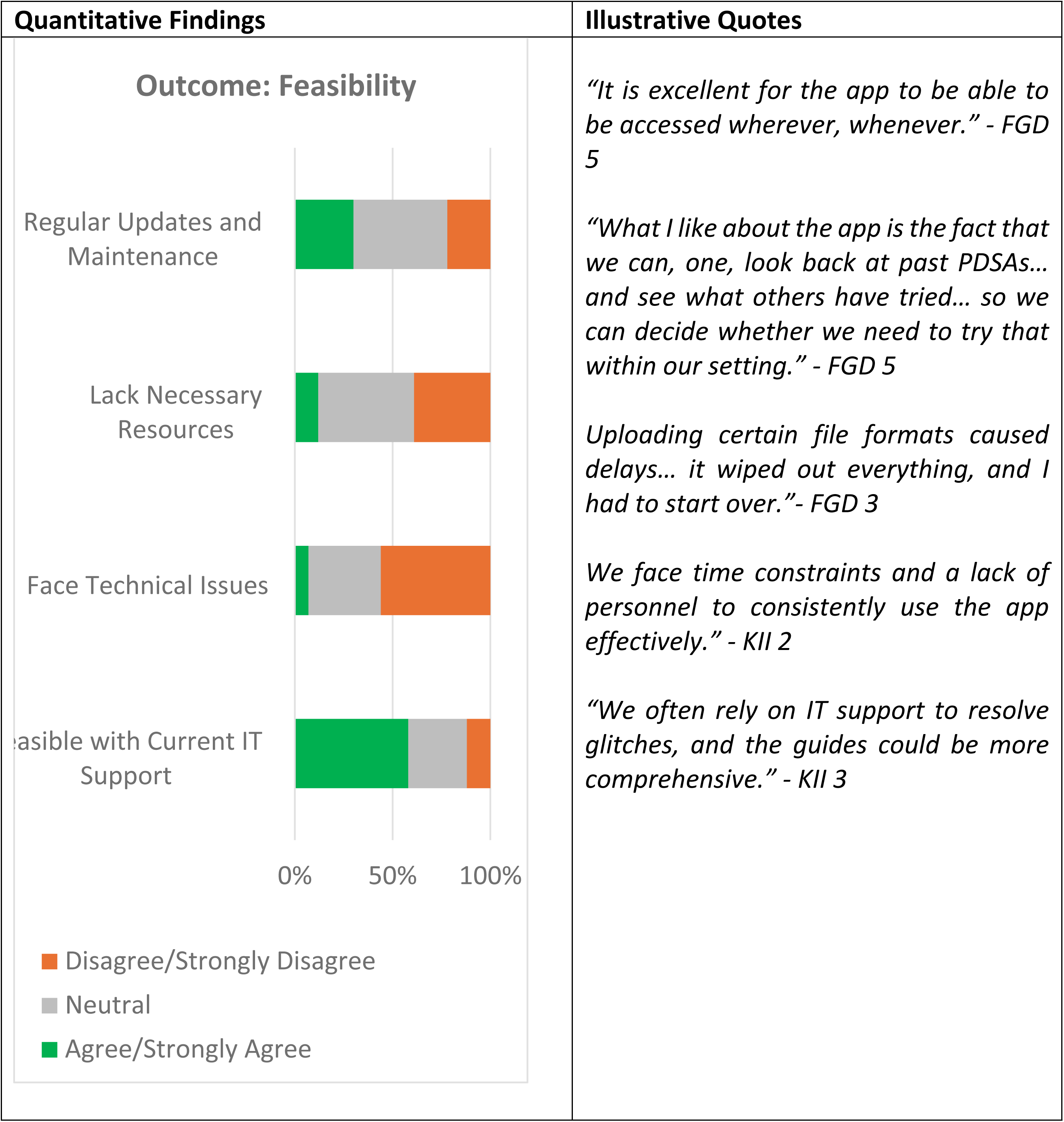
Joint Display of Mixed Methods Findings on the Feasibility of the CQI App. Note: Survey items assessing feasibility included both positive and negatively framed statements. For negatively worded items, disagreement reflects a positive appropriateness outcome.

Participants described several features that facilitated feasibility, including improved documentation processes and the ability to track CQI activities through centralized dashboards and trend visualizations. At the same time, participants reported technical challenges that disrupted routine use, including login difficulties, unstable internet connectivity, and challenges uploading certain file types. Several participants noted that account inactivity sometimes required credential resets, which interrupted continued engagement with the app.

Dependence on facility-level IT support emerged as a recurring theme. While participants generally felt that technical issues could be resolved when escalated, many described limited confidence in troubleshooting problems independently. Participants suggested that embedding user manuals and guides within the app, along with more comprehensive onboarding, could improve self-reliance.

Resource constraints were also frequently cited. Participants described time limitations, staffing shortages, competing responsibilities, and staff turnover as factors that constrained regular updates and sustained use of the app. Suggestions to improve feasibility included ongoing training, clearer expectations for update frequency, and automation of reporting functions to better integrate the CQI app into daily workflows.

## 4. Discussion

This evaluation examined participants’ perceptions of the early-stage implementation of a web-based Continuous Quality Improvement (CQI) platform, the CQI app, within Jamaica’s healthcare system, using four implementation outcomes from the Proctor et al. framework: acceptability, adoption, appropriateness, and feasibility. The evaluation followed substantial groundwork, including participatory adaptation of the CQI app, training workshops, and initial deployment across facilities. Most participants perceived the app as having strong potential to support CQI processes, particularly through improved documentation, visibility of CQI activities, and peer learning. At the same time, participants identified persistent challenges related to infrastructure, continuity of support during rollout, shared accountability for app use, and leadership engagement. As part of formative evaluation, these findings yielded actionable recommendations to strengthen implementation, including clarifying roles and responsibilities for CQI app use, reinforcing leadership ownership, expanding ongoing training and onboarding, and addressing infrastructure and support gaps to enable sustained integration into routine practice.

### Interpretation of Findings in the Context of Jamaica’s Healthcare System

Jamaica has made notable strides in institutionalizing CQI, particularly in HIV care, where collaborative learning and structured quality-improvement processes have strengthened service delivery and improved patient outcomes (14). Within this broader context, participants in this evaluation described varying levels of familiarity and experience with CQI. Nearly half of the survey respondents reported less than one year of experience with CQI frameworks, and many indicated limited use of the CQI app during the early rollout period.

Participants’ limited exposure to CQI and to the CQI app provided important context for interpreting the high proportion of neutral responses observed across several implementation outcomes. During FGDs, participants described still learning how CQI processes fit into their daily work and how the CQI app could support these processes. These findings point to the need to further embed CQI into routine clinical practice, strengthen enabling structures, and deploy digital tools such as the CQI app in ways that support learning, confidence, and sustained use over time (1, 33).

### Leadership and Governance: A Catalyst for CQI and Digital Health Adoption

Strong leadership and governance are crucial to successfully implementing and scaling digital health innovations such as the CQI app (34). This evaluation revealed leadership gaps, as frontline staff lacked the necessary support and guidance to integrate the CQI app into routine healthcare operations. Participants frequently noted that CQI tasks were disproportionately assigned to specific individuals, such as data clerks, creating bottlenecks and undermining team-wide ownership of CQI practices.

The diffusion of innovation theory highlights the role of leadership in fostering an enabling environment that enables early adopters to influence their peers (35). In resource-limited settings like Jamaica, where digital health adoption is still in its infancy, effective leadership is pivotal in driving innovations such as the CQI app (36). Leaders must actively champion these technologies, integrate them into strategic health plans, and align their use with existing facility workflows to ensure sustainability (36). Leadership engagement is crucial for overcoming resistance to change, as many healthcare workers perceive such tools as additional burdens rather than enablers of quality improvement.

Our evaluation underscores the importance of leadership in creating and sustaining an enabling environment for CQI practice as digital tools such as the CQI app are introduced and scaled. Participants’ experiences suggest that digital innovations alone are insufficient to strengthen CQI without foundational systems in place, including clear CQI governance structures, defined roles and responsibilities, standardized reporting processes, and consistent expectations for implementation (37, 38). Leaders play a central role in reinforcing these foundations by promoting shared accountability for CQI activities, supporting standardized approaches to quality improvement, and ensuring access to appropriate tools and resources (1). Within such an enabling environment, the CQI app can serve as an enhancement and accelerator for existing CQI practices, supporting documentation, learning, and the continuity of CQI efforts across the healthcare system.

### IT Infrastructure and Capacity for Digital Health Implementation

These evaluation findings highlight that IT infrastructure remains a critical enabler of digital health implementation in resource-constrained settings. Participants described internet instability, limited device availability, and insufficient IT support as persistent barriers to feasibility. Similar challenges have been documented in other LMIC contexts and in Jamaica [36–38]. Even when digital tools are well designed, routine access issues and platform instability can disrupt integration into daily workflows, undermine user confidence, and discourage sustained use (39).

Participants’ calls for refresher training, onboarding for new staff, and the inclusion of embedded job aids or user manuals underscore the importance of capacity-building as an ongoing process rather than a one-time activity during rollout. These findings align with broader evidence that digital health tools are more likely to succeed when technological functionality is paired with human-centered design and sustained implementation support. Ensuring that healthcare workers, particularly those with limited digital fluency, have access to responsive support systems is critical for bridging the gap between initial adoption and continuous, effective use (33, 36).

### Implications for Practice and Scale-Up

Findings from this early-stage evaluation point to several actionable considerations to strengthen the adoption and scale-up of the CQI app, with broader relevance to digital health implementation.

First, effective integration of digital tools depends on anchoring them within established CQI structures, supported by leadership engagement and clearly defined workflows. Practical entry points include clarifying team roles for CQI documentation, setting expectations for routine app use, and aligning digital tools with existing CQI reporting and supervision processes. Without this alignment, digital tools risk being perceived as add-ons rather than enablers of routine practice.

Second, capacity-building should be approached as a continuous process rather than a one-time training event. Participants’ recommendations highlight the importance of refresher training, onboarding for new staff, and accessible, role-specific support materials. In-built tutorials, job aids, and guided workflows within the CQI app could help address common user barriers, reduce reliance on external support, and strengthen confidence among users with varying levels of digital fluency. Evidence from digital health implementation studies suggests that such embedded supports are critical for sustaining use over time, particularly in resource-constrained settings [30, 33].

Third, strengthening the IT infrastructure remains essential for feasibility and scale-up. Entry points for improvement include ensuring reliable internet connectivity, adequate device availability, and responsive IT support mechanisms. Addressing these foundational constraints can reduce technical disruptions that undermine user confidence and interrupt routine use, even when tools are perceived as valuable.

While participants reported barriers and recommendations specific to the CQI app in Jamaica, their perspectives reflect challenges commonly encountered when implementing digital health tools in similar settings. These findings therefore offer transferable insights for other contexts seeking to digitize quality improvement processes or introduce comparable health information tools.

Finally, this evaluation demonstrates the value of conducting early-stage assessments of implementation outcomes when introducing digital health innovations. By identifying barriers and enablers early, formative evaluations can inform timely course correction, strengthen system readiness, and help prevent digital tools from being underutilized or “withering on the vine” before their potential benefits are realized.

## 5. Strengths and Limitations

This evaluation has several methodological strengths. The use of a convergent mixed-methods design enabled triangulation of survey and qualitative data, strengthening the credibility of findings and allowing for a more nuanced assessment of early implementation experiences. Applying the Implementation Outcomes Framework provided a structured, theory-informed approach for examining acceptability, adoption, appropriateness, and feasibility. Additional strengths include the inclusion of healthcare workers from a broad range of facilities and geographic areas, as well as a relatively high survey response rate for a system-level digital health evaluation conducted during routine service delivery.

However, several limitations warrant consideration. First, the evaluation relied on self-reported data, which introduces the possibility of unknown bias if participants over- or under-reported their experiences or views due to perceived social desirability. At the same time, the survey’s anonymity and the use of group-based qualitative methods may have mitigated this effect by reducing pressure to provide socially acceptable responses.

Second, qualitative participants were selected among healthcare workers attending scheduled CQI learning sessions, which may have led to responses from more engaged or motivated CQI team members. While the repetition of themes across FGDs and KIIs suggested adequate thematic saturation for the evaluation’s formative purpose, the findings may not fully capture the experiences of less-engaged users or facilities with minimal exposure to the CQI app.

Third, the use of both in-person and remote (Zoom-based) data collection modalities may have influenced participant responses, as different interview settings can affect comfort levels and disclosure. Although the same discussion guides and facilitation approach were used across modalities, this variation should be considered when interpreting the qualitative findings.

Finally, this evaluation focused on healthcare worker perceptions and experiences and did not include analysis of system-generated CQI app usage data. As a result, objective measures of adoption and sustained use could not be assessed. In addition, findings reflect the implementation context in Jamaica and may not be generalizable to other settings where the CQI app has been deployed.

Despite these limitations, the evaluation provides a rigorous and contextually grounded assessment of early implementation experiences and offers valuable insights to inform future evaluations and scale-up efforts.

## 6. Conclusion

This evaluation highlights the CQI app’s potential to support quality improvement in Jamaica’s healthcare system while revealing critical enablers for its successful integration. Although the app was generally well-received, its uptake was constrained by infrastructure gaps, limited training, and uneven engagement. Addressing these challenges through strengthened leadership, sustained capacity-building, alignment of foundational CQI systems, and enhanced digital infrastructure is key to scaling its impact. As Jamaica continues to invest in digital health, ensuring an enabling environment will be essential to translating innovation into meaningful and sustainable improvements in the health system. By strengthening CQI documentation and learning in facilities serving populations disproportionately affected by HIV and other chronic conditions, the CQI app has potential equity-relevant benefits in improving consistency and accountability of care delivery.

## Data Availability

The de-identified survey dataset is available upon reasonable request. Requests for access should be directed to The University of Washington, department of Global Health at All other relevant data supporting the findings of this study are included within the manuscript.

## Acknowledgement

This manuscript was supported by the U.S. President’s Emergency Plan for AIDS Relief (PEPFAR) through the Health Resources and Services Administration (HRSA), the U.S. Department of Health and Human Services (HHS) under award number U91HA06801. The award covered 100% of the total costs, totaling $741,495. The contents are those of the author(s). They may not reflect the policies of HRSA, HHS, or the U.S. Government.

